# Cortical scaling of the neonatal brain in typical and altered development

**DOI:** 10.1101/2024.08.15.24311978

**Authors:** Alexandra F. Bonthrone, Daniel Cromb, Andrew Chew, Barat Gal-Er, Christopher Kelly, Shona Falconer, Tomoki Arichi, Kuberan Pushparajah, John Simpson, Mary A. Rutherford, Joseph V. Hajnal, Chiara Nosarti, A. David Edwards, Jonathan O’Muircheartaigh, Serena J. Counsell

## Abstract

Theoretically derived scaling laws capture the non-linear relationships between rapidly expanding brain volume and cortical gyrification across mammalian species and in adult humans. However, the preservation of these laws has not been comprehensively assessed in typical or pathological brain development. Here we assessed the scaling laws governing cortical thickness, surface area and cortical folding in the neonatal brain. We also assessed multivariate morphological terms that capture brain size, shape and folding processes. The sample consisted of 375 typically developing infants, 73 preterm infants and 107 infants with congenital heart disease (CHD) who underwent brain magnetic resonance imaging (MRI). Our results show that typically developing neonates and those with CHD follow the cortical folding scaling law obtained from mammalian brains, children and adults which captures the relationship between exposed surface area, total surface area and cortical thickness. Cortical folding scaling was not affected by gestational age at birth, postmenstrual age at scan, sex or multiple birth in these populations. CHD was characterized by a unique reduction in the multivariate morphological term capturing size, suggesting CHD affects cortical growth overall but not cortical folding processes. In contrast, preterm birth was characterized by altered cortical folding scaling and altered shape, suggesting the developmentally programmed processes of cortical folding are disrupted in this population. The degree of altered shape was associated with cognitive abilities in early childhood in preterm infants.

---

Rapid cortical volumetric expansion and gyrification take place during the third trimester of human pregnancy and early postnatal life (1, 2). The dynamics of cortical development can be captured in two morphological measures, cortical thickness (CT) and surface area (SA), which individually have been found to have distinct spatiotemporal developmental patterns (3, 4), genetic influences (5), and signatures in neurodevelopmental conditions (6–8). However, the development of CT and SA are linked to both volumetric growth and cortical folding processes. Allometric scaling models which employ logarithmic transformation have been used to capture non-linear relationships between anatomical brain characteristics in adults (9, 10) and across mammalian species (11, 12). However, the allometric scaling principles of SA, CT and cortical folding in the typical neonatal brain, and alterations associated with adverse neurodevelopmental conditions, have yet to be fully investigated.

Previous studies investigating the allometric scaling relationships between cortical surface area and brain volume in preterm neonates scanned from birth (25-28 weeks) to 48 weeks PMA report a cortical surface area to supratentorial volume scaling coefficient of 1.27-1.29 (13, 14). Importantly, these studies comprised preterm infants and assessed a wide PMA range, and therefore it is not possible to disentangle typical brain development from the sequelae of preterm birth.

Another universal model describes the scaling laws governing cortical folding in mammalian brains (11) and across children and adult humans (15) by capturing relationships between cortical thickness, total SA, and exposed gyral SA (model details in methods). In this model, changes in one morphological characteristic are necessarily accompanied by compensatory alterations in another, suggesting the dynamics of cortical development may be better captured by multivariate variables capturing the relationships between total SA, exposed SA and CT (16): (i) an offset term characterizing the cellular, molecular and mechanical factors which encourage the cortical surface to fold, (ii) an isometric term capturing size changes, and (iii) a shape term carrying information about shape independent of cortical size or folding (15, 16).

To our knowledge no study has comprehensively characterized allometric scaling or multivariate morphometric terms in the typical neonatal brain or how they may be altered in altered neurodevelopmental conditions at around the time of typical delivery (≥37 PMA). The first aim of this study was to characterize the allometric scaling coefficients governing total SA, CT and cortical folding as well as multivariate morphological terms (offset, isometric and shape) and associations with sex, gestational age at birth (GA), PMA, and multiple birth in the typical neonatal brain using MRI data collected from the developing human connectome project (dHCP) (17). The second aim was to assess how exposure to adverse developmental conditions alters cortical scaling or multivariate morphology, focusing on two distinct populations: infants born preterm (infants in dHCP born <33+0 weeks GA) and infants with congenital heart disease (CHD) requiring surgery within the first year after birth (imaged using the same scanner and acquisitions as dHCP (18)). The final aim was to investigate if allometric scaling or multivariate morphology are associated with early childhood neurodevelopmental outcomes in typical and clinical populations.

## Results

T2-weighted neonatal brain MR images were acquired at 37-44+6 weeks PMA from 345 healthy control infants delivered at term, 73 preterm infants and 107 infants with CHD without major lesions (table S1 contains demographics). Images were processed using the dHCP neonatal cortical surface reconstruction pipeline (19) to provide measures of total SA, exposed SA, mean curvature corrected CT and supratentorial brain volume. Analyses were conducted in R v.4.4.0 using linear regressions with permutation testing (n=5000).

### Scaling coefficients in typical and altered development

In typically developing infants, the cortical folding scaling coefficient was not significantly different from the cross-species coefficient of 1.25 (α [95% CI] 1.26 [1.21-1.30], p=0.719). The total SA scaling coefficient (0.873 [0.830-0.914]) was significantly different from the theoretical coefficient of 2/3 but did not differ significantly from the coefficient previously observed in a normative adult sample (α=0.89, p=0.418)(9). In contrast, the CT scaling coefficient (0.087 [0.040-0.132]) significantly differed from both the theoretical (1/3, p<0.001) and previously observed adult (0.03, p=0.017) coefficients (9).

The preterm allometric scaling coefficient for cortical folding (α [95% CI] 1.41 [1.30-1.51]) was significantly different from controls and infants with CHD (Table 1; Figure 1a), as well as the cross-species coefficient (p=0.004). The cortical folding scaling coefficient in CHD (1.18 [1.08-1.28]) was not significantly different from controls (Table 1), or the cross-species coefficient (p=0.165).

**Table 1.** The effect of group on cortical allometric scaling relationships.

|  |  |  |  |
| --- | --- | --- | --- |
| Model | B (95% CI) | T | P (p<sub>FWE</sub>) |
| **Typically developing controls (reference) and preterm infants** |  |  |  |
| *Allometric scaling coefficients* |  |  |  |
| Log(SA)~Log(supratentorial volume) | 0.029 (-0.044-0.103) | 0.782 | 0.447 (0.894) |
| Log(CT)~ log(supratentorial volume) | 0.012 (-0.059-0.083) | 0.331 | 0.749 (0.894) |
| Log(total SA*CT<sup>1/2</sup>) ~ log(exposed SA) | 0.128 (0.033-0.222) | 2.65 | 0.011 (0.033)* |
| *Multivariate morphological terms* |  |  |  |
| Offset | -0.013 (-0.019- -0.008) | -4.80 | <0.001 (<0.001)* |
| Isometric | 0.020 (-0.010- 0.050) | 1.30 | 0.204 (0.204) |
| Shape | -0.140 (-0.188- -0.092) | -5.71 | <0.001 (<0.001)* |
| **Typically developing controls (reference) and infants with CHD** |  |  |  |
| *Allometric scaling coefficients* |  |  |  |
| Log(SA)~Log(supratentorial volume) | -0.011 (-0.088-0.065) | -0.295 | 0.243 (0.820) |
| Log(CT)~ log(supratentorial volume) | -0.036 (-0.114- 0.041) | -0.928 | 0.772 (0.820) |
| Log(total SA*CT<sup>1/2</sup>) ~ log(exposed SA) | 0.094 (-0.024-0.212) | 1.57 | 0.250 (0.820) |
| *Multivariate morphological terms* |  |  |  |
| offset | -0.003 (-0.008-0.002) | -1.11 | 0.205 (0.205) |
| isometric | -0.034 (-0.061- -0.006) | -2.43 | 0.003 (0.008)* |
| shape | -0.056 (-0.103- -0.009) | -2.36 | 0.046 (0.091) |
| **preterm infants (reference) and infants with CHD** |  |  |  |
| *Allometric scaling coefficients* |  |  |  |
| Log(SA)~Log(supratentorial volume) | -0.018 (-0.108- 0.072) | -0.387 | 0.701 (0.701) |
| Log(CT)~ log(supratentorial volume) | -0.073 (-0.164- 0.018) | -1.58 | 0.110 (0.220) |
| Log(SA*CT<sup>1/2</sup>) ~ log(exposed SA) | -0.142 (-0.252- -0.032) | -2.54 | 0.010 (0.030)* |
| *Multivariate morphological terms* |  |  |  |
| offset | 0.013 (0.007- 0.020) | 4.31 | <0.001 (<0.001)* |
| isometric | -0.035 (-0.077- 0.006) | -1.67 | 0.099 (0.099) |
| shape | 0.081 (0.013- 0.149) | 2.35 | 0.018 (0.036)* |
| * significant $p_{\text{FWE}} < 0.05$; model covariates included PMA, PMA<sup>2</sup>, sex and multiple birth. GA also included in model with CHD and typically developing controls |  |  |  |

**Figure 1.**
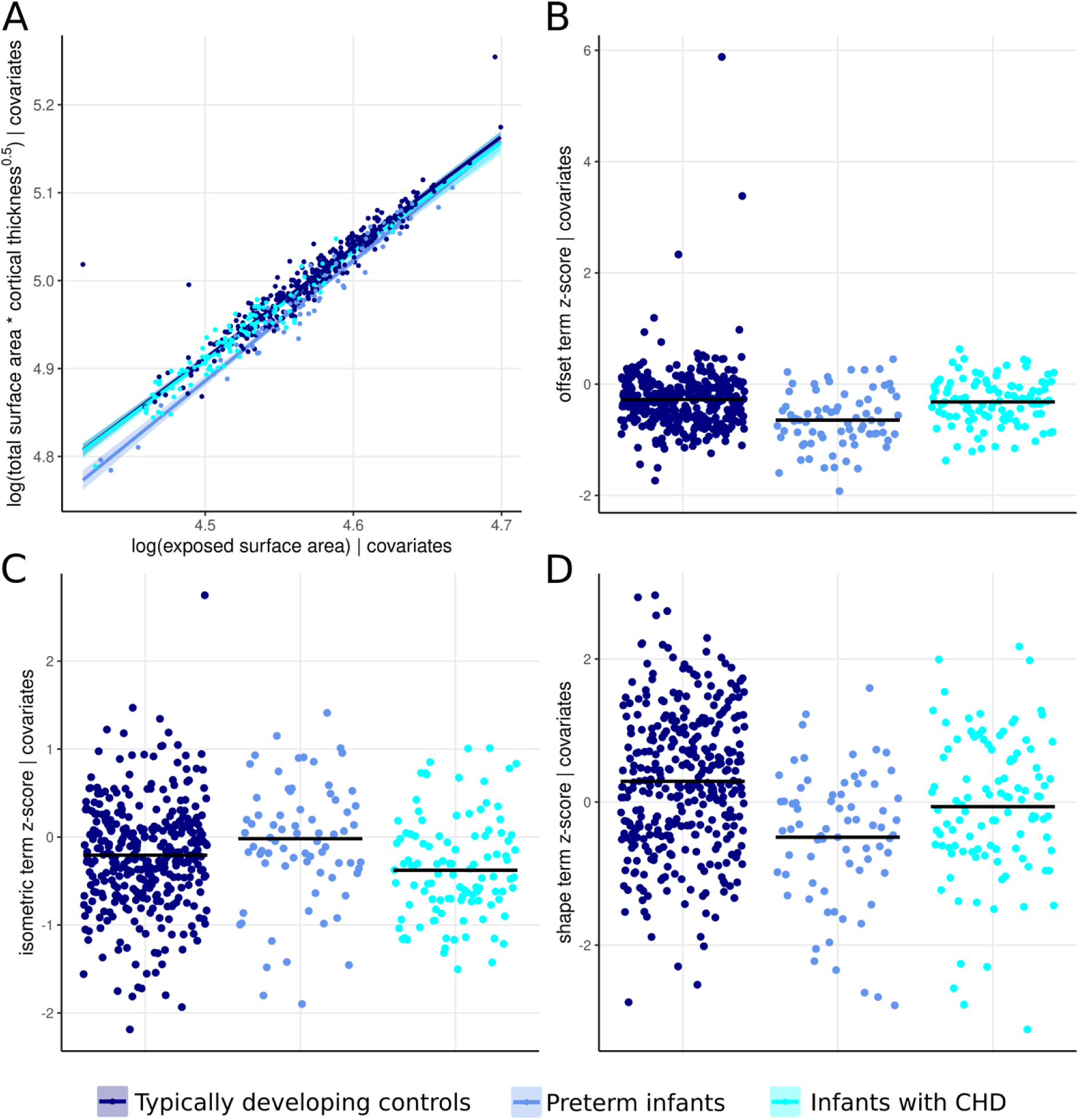
Differences in A) folding scaling, and multivariate cortical terms (B-D) between groups adjusting for PMA, PMA^2^, sex and multiple birth. Variables in B-D were z-scored to controls for visualization purposes as terms are unitless.

The allometric scaling coefficients for total SA (α [95% CI] preterm=0.893 [0.820-0.968], CHD=0.855 [0.770-0.940]) and CT (preterm=0.079 [0.020-0.138], CHD=0.095 [0.090-0.171]) were not significantly different from controls (Table 1), but significantly differed from the theoretical coefficients (all p<0.001). There were no significant associations between demographics and scaling coefficients in any group (p_FWE_>0.05, tables S2 and S3).

### Multivariate morphology in typical and altered development

In controls, GA was significantly associated with multivariate morphology (offset B [95% CI] - 0.003 (-0.006-<-0.001), t=2.66, p_FWE_=0.049; isometric -0.037 [-0.048--0.025], t=-5.99, p_FWE_=<0.001; shape 0.041 [0.020-0.061], t=3.87, p_FWE_<0.001) (figure S1). Multiple birth was also associated with the isometric term (B [95% CI] -0.090 [-0.155--0.024], t=0.20, p_FWE_=0.024) (figure S1).

When compared to both controls and infants with CHD, preterm infants had significantly altered offset and shape terms (Table 1; Figure 1b and c). Infants with CHD had lower isometric terms compared to controls (Table 1; Figure 1d). In preterm infants, the degree of prematurity (i.e. GA at birth) was associated with offset (B [95% CI] 0.003 [0.002-0.004], t=4.10, p_FWE_=0.006) and isometric (0.016 [0.007-0.025], t=3.39, p_FWE_=0.043) terms. The isometric term was also associated with PMA (1.10 [0.486-1.71], t=0.3.57, p_FWE_=0.017) and PMA^2^ (-0.012 [-0.020--0.005], t=-3.31, p_FWE_=0.012) in preterm infants. There were no other significant relationships between multivariate morphology and demographics in any groups (p_FWE_>0.05, table S2, table S3).

### Multivariate shape is associated with early childhood cognitive abilities in preterm infants

All controls, 57 preterm infants and 65 infants with CHD underwent a Bayley scales of infant and toddler development-3^rd^ edition (20) assessment at 17.26-40.29 months corrected age providing cognitive, language and motor composite scores.

An individual’s degree of deviation from the typical population across each scaling metric and multivariate morphological term was calculated as previously reported (9). A complete linear model for each metric was constructed for the typically developing control population and then applied to infants with CHD and preterm infants. Residuals were extracted and z-scored to the controls to measure an individual’s deviance from the typical population, given their supratentorial volume/exposed SA (for scaling relationships), PMA, PMA^2^, sex and multiple birth status.

There were no significant associations between deviance z-scores and neurodevelopmental outcomes in typically developing infants (p_FWE_>0.05, table S4). There was a significant interaction between group and shape term deviance z-score when predicting cognitive composite scores (Table 2; Figure 2b). The z-score predicted cognitive scores in preterm infants (p=0.009, bootstrapped R^2^ for shape z-score=0.175), but not controls (p=0.250) or CHD (p=0.228). Post-hoc analysis revealed the interaction remained significant when excluding controls (p=0.004). There were no other significant interactions between deviance z-scores and neurodevelopmenatal outcomes (Table 2).

**Figure 2.**
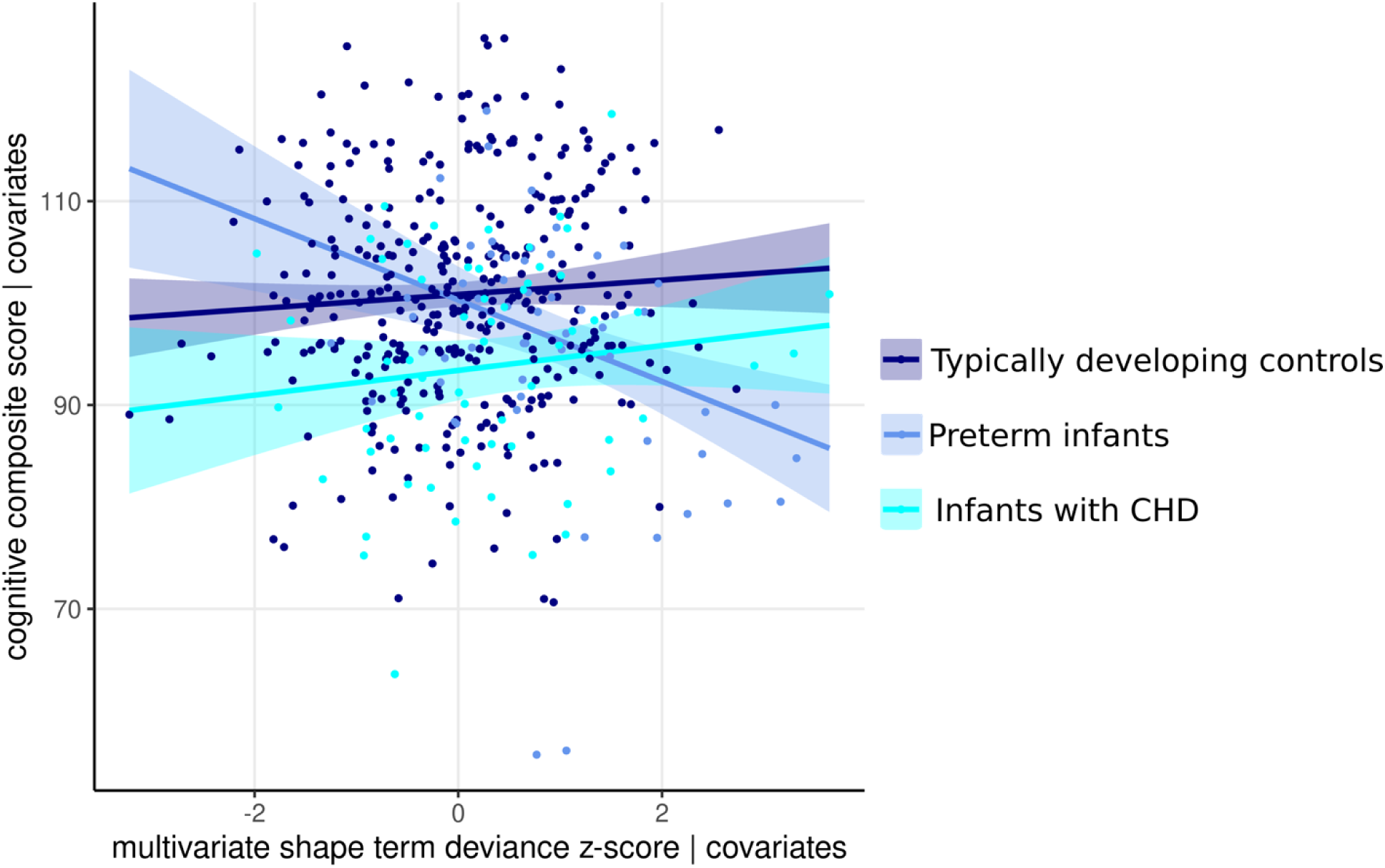
Relationship between multivariate shape term deviance z-score and cognitive composite score across each group adjusting form socioeconomic status

**Table 2.** Interactions between group and scaling relationship/multivariate morphological deviance z-scores when predicting outcomes.

| Model | B (95% CI) | T | p |
| --- | --- | --- | --- |
| **Cognitive composite scores** |  |  |  |
| *Preterm infants* |  |  |  |
| Log(total SA) | -2.84 (-6.02- 0.343) | -1.75 | 0.084 (0.933) |
| Log(mean CT) | 4.17 (1.06- 7.28) | 2.63 | 0.009 (0.120) |
| Log(total SA*mean CT<sup>1/2</sup>) | -3.88 (-7.49- -0.271) | -2.10 | 0.037 (0.480) |
| Offset | -0.776 (-4.13- 2.57) | -0.455 | 0.641 (1.00) |
| isometric | -0.156 (-3.00- 2.69) | -0.108 | 0.919 (1.00) |
| shape | -4.78 (-7.84- -1.73) | -3.08 | 0.002 (0.040)* |
| *Infants with CHD* |  |  |  |
| Log(total SA) | -1.27 (-4.16- 1.63) | -0.681 | 0.401 (1.00) |
| Log(mean CT) | -1.29 (-4.02- 1.45) | -0.923 | 0.357 (1.00) |
| Log(total SA*mean CT<sup>1/2</sup>) | -2.44 (-5.82- 0.943) | -1.42 | 0.153 (1.00) |
| offset | -4.92 (-891- -0.927) | 2.42 | 0.018 (0.324) |
| isometric | -2.08 (-5.19 - 1.02) | -1.32 | 0.191 (1.00) |
| shape | 0.797 (-1.87- 3.46) | 0.587 | 0.560 (1.00) |
| **Language composite scores** |  |  |  |
| *Preterm infants* |  |  |  |
| Log(total SA) | -3.18 (-7.61- 1.25) | -1.41 | 0.160 (1.00) |
| Log(mean CT) | 6.31 (1.71- 10.9) | 2.70 | 0.007 (0.112) |
| Log(total SA*mean CT<sup>1/2</sup>) | -5.17 (-10.2- -0.155) | -2.03 | 0.050 (0.602) |
| offset | -0.651 (-5.46- 4.16) | -0.266 | 0.792 (1.00) |
| isometric | 2.36 (-1.94- 6.65) | 1.08 | 0.285 (1.00) |
| shape | -6.06 (-10.5- -1.64) | -2.70 | 0.007 (0.112) |
| *Infants with CHD* |  |  |  |
| Log(total SA) | -2.87 (-7.43- 1.68) | -1.24 | 0.218 (1.00) |
| Log(mean CT) | -1.52 (-5.81- 2.76) | -0.697 | 0.500 (1.00) |
| Log(total SA*mean CT<sup>1/2</sup>) | -3.59 (-8.80- 1.61) | -1.36 | 0.175 (1.00) |
| offset | -5.77 (-11.7- 0.111) | -1.93 | 0.053 (0.952) |
| isometric | -2.16 (-6.99- 2.66) | -0.881 | 0.375 (1.00) |
| shape | 0.938 (-3.22- 5.10) | 0.443 | 0.663 (1.00) |
| **Motor composite scores** |  |  |  |
| *Preterm infants* |  |  |  |
| Log(total SA) | -1.76 (-4.68- 1.15) | -1.19 | 0.239 (1.00) |
| Log(mean CT) | 1.68 (-1.18- 4.54) | 1.15 | 0.248 (1.00) |
| Log(total SA*mean CT<sup>1/2</sup>) | -4.72 (-8.01- -1.43) | -2.82 | 0.006 (0.099) |
| offset | -2.63 (-5.70- 0.442) | -1.68 | 0.095 (0.952) |
| isometric | 0.407 (-2.19- 3.00) | 0.308 | 0.757 (1.00) |
| shape | -1.82 (-4.63- 0.992) | -1.27 | 0.211 (1.00) |
| *Infants with CHD* |  |  |  |
| Log(total SA) | -0.272 (-2.94- 2.40) | -0.200 | 0.845 (1.00) |
| Log(mean CT) | -0.501 (-3.02- 2.02) | -0.390 | 0.690 (1.00) |
| Log(total SA*mean CT<sup>1/2</sup>) | -0.593 (-3.68- 2.50) | -0.377 | 0.707 (1.00) |
| offset | -1.59 (-5.25- 2.07) | -0.855 | 0.398 (1.00) |
| isometric | 0.581 (-3.42- 2.26) | -0.402 | 0.672 (1.00) |
| shape | 0.393 (-2.07- 2.86) | 0.314 | 0.760 (1.00) |
| *significant results $p_{FWE} < 0.05$; linear regression with 5000 permutations, model covariate included index of multiple deprivation. Additional covariate for language models was if a parent's first language was not english | | | |

### Assessing individual cortical metrics

The effect of group on total SA, CT, gyrification index (total SA/exposed SA; GI) and supratentorial brain volume was assessed (table S5). Preterm infants had significantly lower SA, GI and supratentorial volume, and higher CT compared to controls (p_FWE_≤0.007). Infants with CHD had lower total cortical SA, GI, and supratentorial brain volume (p_FWE_≤0.010). GI was significantly lower in infants with CHD compared to preterm infants (p_FWE_<0.001). There were no associations between outcome measures and individual cortical metrics (pFWE>0.05, table S6).

## Discussion

In this study, we demonstrate that typical neonatal cortical development conforms to the intrinsic principle folding scaling law that applies to child and adult human and other mammalian brains. Following preterm birth, but not CHD, the folding scaling coefficient was significantly different from the control population. Preterm birth was also characterized by reduced multivariate morphological shape and offset terms, while CHD was characterized by reduced isometric terms with intact offset and shape characteristics. These data suggest distinct cortical developmental changes in different at-risk populations. Scaling coefficients and multivariate morphology were not associated with neurodevelopmental outcomes in typical infants or those with CHD. In contrast, for a given preterm infant, the shape term’s degree of deviation from the typical population was associated with later cognition in early childhood. Finally, there were no differences in scaling principles of total SA and CT in the clinical cohorts.

The typically developing neonatal brain follows the same cortical folding scaling law as typical child and adult brains (15) and other mammalian cortices (11). The folding scaling coefficient may therefore represent information about the molecular, cellular and mechanical mechanisms that shape mammalian cortical folding (21). The allometric scaling coefficient linking SA and supratentorial volume in typically developing neonates is significantly higher than the theoretical geometric scaling coefficient of 0.67, but not significantly different from the coefficient previously reported in the typical adult population (9). These results suggest that in the brains of typical infants born around the expected date of delivery (≥37 weeks), as in the typical adult brain (22), higher volume is accompanied by a larger increase in SA, reflecting increased gyrification. The scaling coefficient between CT and volume, while significantly lower than the theoretical coefficient, was significantly higher than that reported in typical adults (9). In typical brain development, CT increases rapidly to a peak around 2 years before declining across adolescence and adulthood, while cerebral volume and surface area increase into adolescence (4), which may explain the difference in scaling relationships. Together, our results suggest that, at the time of normal delivery, the typically developing brain follows the same scaling relationships for cortical folding and SA as adults which may reflect biologically conserved folding mechanisms. In contrast, the scaling coefficient for CT in infants was significantly higher than in adults, perhaps reflecting the unique developmental trajectory of CT in early childhood.

In preterm infants, the scaling coefficient for cortical folding is significantly different from the corresponding value in typically developing controls and the theoretically predicted value. The multivariate morphological term capturing factors encouraging folding (offset) was also significantly reduced in preterm infants compared to controls. The neurobiological mechanisms of altered cortical folding in premature infants are unclear. Reduced folding may be related to altered underlying white matter connections in preterm infants (23, 24). The scaling results suggest that altered cortical development following preterm birth in the absence of major destructive lesions is not due to fewer cortical neurons (11, 25). We identified lower supratentorial brain volume, total SA and GI but thicker cortex in preterm infants compared to controls, a finding supported by previous studies (6, 26). Additionally, histological research suggests preterm birth alters neuronal maturation but is not associated with large-scale neuronal loss (27). Furthermore, adults born preterm show altered cortical folding scaling coefficients and multivariate morphological offset terms compared to controls (28), suggesting preterm birth alters developmentally programmed folding principles with effects across the lifespan.

We also find the multivariate morphological shape term is altered in infants born preterm and that this term predicts cognitive abilities in later childhood. These findings complement previous research in preterm infants which has identified associations between cognition in early childhood and regional changes in shape characteristics such as CT (29), sulcal depth (30) and cortical curvature (31) at the same timepoint as our study (37-44+6 weeks PMA). These effects may have long lasting implications, as total SA growth from 22-44 weeks PMA has also been found to be associated with cognitive abilities in middle childhood in preterm infants (7).

Two previous studies reported the scaling coefficient between total cortical SA and volume in preterm infants was 1.27-1.29, significantly higher than the values reported in our study (13, 14). However, it is important to note that infants in previous studies were scanned from birth (22-30 weeks) to 45-48 weeks PMA, whereas our analyses were restricted to infants scanned at term equivalent age only. A small study comparing infants born preterm to fetuses imaged from 21 to 36 weeks GA suggests allometric scaling between SA and brain volume is significantly higher in preterm infants, however direct comparison between fetal and neonatal MR data remains challenging (32). This may reflect a different developmental period, as gyrification increases rapidly from around 28 weeks GA (1, 2). Kapellou and colleagues previously reported lower GA and male sex are associated larger alterations in cortical SA scaling in preterm infants (13), which we did not replicate. The effect of sex and dose-dependent effects of prematurity may be stronger earlier in cortical development. Future research across the fetal and early preterm period is required to fully characterize allometric scaling across development.

In contrast to preterm infants, infants with CHD had significantly lower multivariate morphometric isometric terms than controls but showed no significant differences in scaling coefficients, offset or shape terms. CHD is associated with chronic suboptimal supply of oxygen, glucose and other nutrients that support optimal fetal and neonatal brain development (33–35). In keeping with this, several studies have identified reduced brain volumes in fetuses and neonates with CHD compared with healthy infants (36–39) the degree of which was associated with reduced cerebral substrate delivery (18, 33 39). Reduced cortical folding has also been reported in infants with CHD (18, 40, 41), however Cromb, Wilson and colleagues (42) demonstrated that differences in GI between infants with CHD and controls were explained by supratentorial brain volume rather than a specific deficit in folding. Here we support these findings by demonstrating that the allometric scaling of cortical folding is not significantly different from either typically developing infants or the cross-species coefficient (11). The primary cerebral phenotype in CHD may therefore be smaller brain volumes, with proportionate reductions in cortical folding, rather than specifically disrupted folding mechanisms.

Importantly, when assessing individual cortical metrics, it is possible to conclude that infants with CHD and preterm infants have similar cerebral changes, reflected in reductions in cortical volume, SA, and GI compared to controls. However, allometric scaling relationships and multivariate morphology demonstrate distinct alterations, with CHD showing reduced brain size while preterm infants show aberrant cortical folding and shape compared to controls. Further evidence that the differences in brain development and distinct in CHD and preterm infants can be seen in the differing patterns of cortical microstructural changes reported in these groups, with preterm infants displaying widespread reductions in neurite density index and increases in mean diffusivity (6), while infants with CHD show reductions in fractional anisotropy and orientation dispersion index and no differences in neurite density or mean diffusivity (43). Our work highlights the value of considering multiple covarying morphological variables when distinguishing phenotypes between clinical groups.

Our results have some limitations which are important to acknowledge. We used whole brain measures to assess cortical morphology; however, there are likely regional variations in scaling behaviors in typically developing infants and clinical groups. In addition, our analysis assessed infants from 37+0-44+6 PMA, however the dynamics of cortical scaling are likely different across different development periods. Future research should assess morphological development across the fetal and preterm period.

In summary, the typical neonatal brain conforms to the scaling law governing cortical folding in children, adults and other mammalian brains. We report distinct cortical developmental changes in different at-risk populations. Brains were smaller in infants with CHD, while preterm infants demonstrated altered cortical folding and brain shape compared to controls. Shape characteristics were associated with early childhood cognition in preterm infants but not controls or CHD.

## Materials and Methods

### Ethical approval

The National Research Ethics Service provided ethical approval (CHD: 07/H0707/105, 21/WA/0075; dHCP 14/LO/1169). In accordance with the declaration of Helsinki, informed written parental consent was obtained before neonatal MRI and neurodevelopmental assessment.

### Recruitment

Data from controls and preterm infants were from the dHCP 3^rd^ neonatal data release (17). The control sample included 345 infants. Exclusion criteria were GA <37 weeks, admission to the neonatal intensive care unit, a 1st degree relative with a diagnosed neurodevelopmental condition (44), a neurodevelopmental outcome score below 70 (<2 standard deviations from the test mean) and no evidence of walking at neurodevelopmental assessment.

The preterm sample consisted of 73 infants born 23-32+6 weeks GA. Exclusion criteria included no brain MRI at ≥37 weeks PMA and suspected/confirmed chromosomal abnormality/congenital syndrome.

Infants with CHD requiring cardiac catheterization or surgery before one-year postnatal age (45) were recruited as fetuses or neonates from St Thomas’ Hospital, London October 2015-July 2023. Exclusion criteria were birth <37 weeks GA, suspected/confirmed chromosomal abnormality/congenital syndrome, previous neonatal surgery (excluding cardiac catheterization) or suspected congenital infection (18).

Exclusion criteria for all infants were no/motion corrupted neonatal brain MRI and major lesions on MRI (e.g. arterial ischemic stroke or parenchymal haemorrhage) after review by two perinatal neuroradiologists.

### MRI acquisition

Brain MRI was performed during natural sleep following feeding on a Philips Achieva (Best, Netherlands) 3 Tesla system situated in the neonatal intensive care unit at St. Thomas’ Hospital using a 32-channel neonatal head coil and neonatal positioning device (Rapid Biomedical GmbH, Rimpar DE) (46). Hearing protection included silicone-based putty earplugs placed in the external auditory meatus (President Putty), neonatal earmuffs (MiniMuffs) and an acoustic hood placed over the infant. A pediatrician or nurse trained in MR procedures supervised scanning and monitored pulse oximetry, axillary temperature, electrocardiography, and respiratory rate throughout.

T2-weighted multi-slice turbo spin echo scans were acquired in sagittal and axial planes [repetition time (TR)/echo time (TE) 12,000/156 ms; flip angle 90°; slice thickness 1.6 mm; slice overlap 0.8 mm; in-plane resolution 0.8×0.8 mm; SENSE factor 2.11/2.58 (axial/sagittal)] with a 5s noise ramp-up to avoid a startle response. T2-weighted volumes were reconstructed using a dedicated algorithm (reconstructed voxel size = 0.5 mm^3^) (47).

### MRI processing

T2-weighted brain MRI were processed using the dHCP structural pipeline (19). Reconstructed T2-weighted images were corrected for bias-field inhomogeneities, brain extracted and segmented into nine tissue classes using the Draw-EM algorithm (48). Supratentorial brain volume was calculated by summing cortical white matter, cortical grey matter, hippocampus and amygdala and deep grey matter volumes. Left and right white, pial and mid-thickness surfaces were extracted and inflated. CT (curvature regressed out) and total pial SA were calculated and extracted excluding the midline. The area of the exposed pial cortex was calculated using the medical image registration toolkit (https://mirtk.github.io/) and connectome workbench v.1.5.0.

### Neurodevelopmental assessment

Infants were invited to attend a neurodevelopmental follow-up assessment as close to 18 months (dHCP) or 22 months (CHD) corrected age as possible. The Bayley Scales of Infant and Toddler Development-3^rd^ Edition (20) were administered by a developmental paediatrician or psychologist to obtain cognitive, language and motor composite scores [test mean (SD) 100(15)]. In addition, parents completed a questionnaire which asked whether English was the first language of the child’s parents.

### Socioeconomic status

IMD was calculated using the 2019 data release for postcode at birth and reported as ranks and quintiles (most to least deprived). IMD is a composite measure of socioeconomic status in England encompassing factors including income, employment, education, health and crime.

### Statistical analysis

All analyses were conducted in R v.4.4.0. All demographic variables were presented with median and inter-quartile ranges.

#### Allometric Scaling

The geometric scaling laws governing surface thickness, area and total volume in three-dimensional objects of identical shape but different size are as follows:

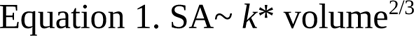

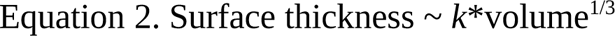

In these equations, *k* represents a constant and 2/3 and 1/3 represent the scaling exponents (*α).* By logarithmically transforming these equations, *α* for each cortex was measured as the regression coefficient (B) in a linear model:

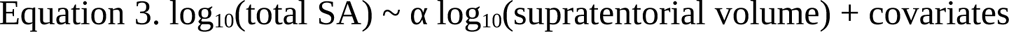

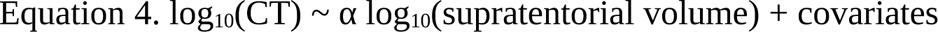

The universal scaling model governing cortical folding is described in equation 5, where α=1.25, and *α* was calculated for all infants adjusting for covariates (Equation 6).

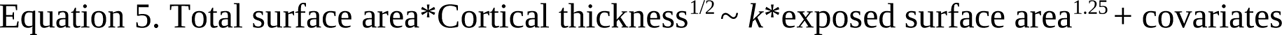

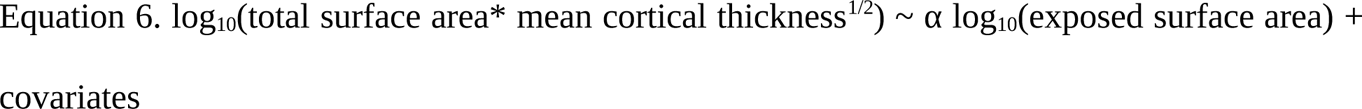

Multiple linear regressions were used to estimate scaling coefficients in controls, preterm infants and infants born prematurely adjusting for PMA, PMA^2^, sex and multiple birth. Scaling coefficients with 95% confidence intervals were calculated from the regression coefficient and standard error for log(supratentorial volume) or log(exposed SA). The GVLMA package (https://cran.r-project.org/web/packages/gvlma) was used to check model fit and if models did not meet regression assumptions, robust regression with fast-S algorithms were used to estimate regression coefficients (49).

#### Multivariate morphological terms

Geometrically, each cortex can be represented by a point across three axes {total surface area, exposed surface area, thickness^2^}(16) and the scaling law in equation 6 represents the plane close to which cortices are expected to plot. The deviation from this plane was captured by isolating K, the statistical offset:

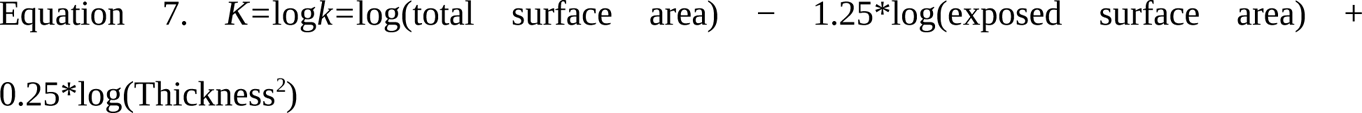

An isometric term was calculated by multiplying all morphological variables by a common numerical factor (e.g. 1) to capture size changes:

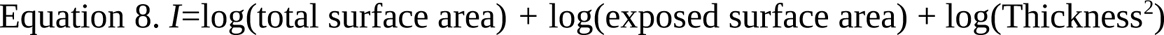

A final shape variable, the cross product of coefficients from offset {1, -1.25,0.25} and isometric terms {1,1,1}, was calculate to carry information about shape, independent of cortical size or the offset.

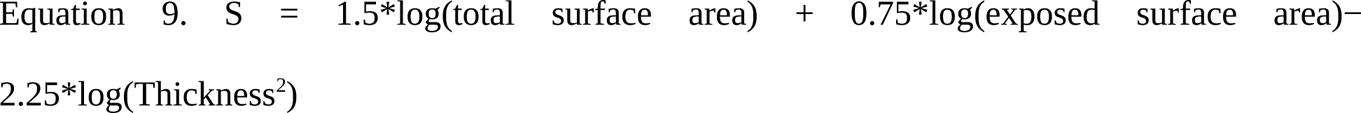

#### Statistical comparisons

The linearHypothesis function in the CAR package (https://r-forge.r-project.org/projects/car/) was used to test if observed regression coefficients were significantly different than the theoretically predicted values and observed coefficients in adults (9).

Multiple linear regressions with permutation testing (n=5000) were used for all further analyses. The effect of GA, PMA, PMA^2^, sex and multiple birth on scaling coefficients and multivariate morphological terms in each group was assessed. To assess the effect on scaling coefficients, the interaction between each variable and log(supratentorial volume) or log(total exposed surface area) was assessed. For multivariate morphological terms, all variables were entered into the models as independent predictors.

To assess the effect of group on scaling coefficients, the interaction between group and log(supratentorial volume) or log(total exposed surface area) was assessed. For multivariate morphological terms, group was entered into the models as an independent predictor. Analyses were undertaken separately to assess the effect of group in preterm infants and controls, and preterm infants and infants with CHD adjusting for PMA, PMA^2^, sex and multiple birth. To assess the effect of group in infants with CHD and controls, typically developing infants who underwent MRI outside of the PMA range of infants with CHD (37.14-42.29 weeks) were removed (n=112). The effect of group (control vs CHD) was assessed adjusting for PMA, PMA^2^, GA, sex and multiple birth.

In order to characterize the association between scaling relationships, multivariate morphological terms and neurodevelopmental outcomes in early childhood, an individual’s degree of deviation from the typical population across each metric was calculated as previously reported by Williams and colleagues (9). A complete robust regression model for each metric was constructed for the typically developing control population and then applied to infants with CHD and those born prematurely. For each metric, residuals were extracted for all infants, and z-scored to the control residuals to obtain a z-score measuring an individual’s deviance from the typical population, given their PMA, PMA^2^, sex, multiple birth status and additionally log(supratentorial volume) or log(exposed SA) for scaling relationships. We assessed if deviance z-scores predicted cognitive, language and motor scores in early childhood in typically developing controls, and if there was an interaction between group (typically developing control, preterm, CHD) and deviance z-scores adjusting for IMD. We also covaried for a parent speaking English as a second language in the language models. Bootstrapped R^2^ were calculated for each deviance z-score which predicted neurodevelopmental outcome scores.

Holm family-wise error rate correction was applied to adjust for multiple comparisons (reported as p_FWE_).

## Data availability

Data from the dHCP included in this study is from the 3^rd^ neonatal data release (https://www.developingconnectome.org/)(18). Processed data from infants with CHD are available from the corresponding author upon reasonable request.

## Acknowledgments

This research was funded by the Medical Research Council (MRC) UK [MR/L011530/1; MR/V002465/1], the British Heart Foundation [FS/15/55/31649] and Action Medical Research [GN2630]. The Developing Human Connectome Project was funded by the European Research Council under the European Union’s Seventh Framework Program [FP7/20072013]/European Research Council grant agreement no. 319456. This research was supported by the Wellcome Engineering and Physical Sciences Research Council Centre for Medical Engineering at King’s College London [WT 203148/Z/16/Z], and by the National Institute for Health Research (NIHR) Biomedical Research Centre based at Guy’s and St Thomas’ NHS Foundation Trust and King’s College London. JO is supported by a Sir Henry Dale Fellowship jointly funded by the Wellcome Trust and the Royal Society [206675/Z/17/Z]. TA receives funding from the MRC via a translation support award [MR/V036874/1] and a senior clinical fellowship [MR/Y009665/1]. ADE, JO, MAR and TA receive funding from the MRC Centre for Neurodevelopmental Disorders, King’s College London [MR/N026063/1]. The views expressed are those of the authors and not necessarily those of the NHS, the NIHR or the Department of Health.

We would like to thank the families who participated in this study. We also thank our research radiologists, our research radiographers, and our neonatal scanning team. In addition, we thank the staff from the St Thomas’ Hospital Neonatal Intensive Care Unit; the Evelina London Children’s Hospital Fetal and Paediatric Cardiology Departments; the Evelina London Paediatric Intensive Care Unit; and the Centre for the Developing Brain at King’s College London.

## Competing interests

The authors declare no competing interests.

## Notes

### Competing Interest Statement

The authors have declared no competing interest.

### Author Declarations

The National Research Ethics Service provided ethical approval (CHD: 07/H0707/105, 21/WA/0075; dHCP 14/LO/1169).

