## Supplementary material for "Cortical scaling of the neonatal brain in typical and altered development": Table S1-S6; Figure S1

| **Table S1.** Sample characteristics | | | |
| --- | --- | --- | --- |
|  | Controls (n=345) | Preterms (n=73) | CHD (n=107) |
| GA at Birth, median (IQR) | 40.14 (39.00-40.86) | 29.14 (27.43-31.29) | 38.57 (38.07-38.93) |
| PMA at Scan, median (IQR) | 41.43 (40.29-42.86) | 41.29 (40.00-42.86) | 39.00 (38.57-39.71) |
| Male, n (%) | 179 (51.9) | 39 (53.4) | 60 (56.1) |
| Index of Multiple Deprivation Quintile | | | |
| 1 (most deprived) | 52 (15.1) | 13 (17.8) | 17 (15.9) |
| 2 | 138 (40.0) | 23 (31.5) | 18 (16.8) |
| 3 | 70 (20.3) | 19 (26.0) | 26 (23.4) |
| 4 | 32 (9.3) | 4 (5.5) | 19 (17.8) |
| 5 | 51 (14.8) | 14 (19.2) | 26 (24.3) |
| Not Available | 2 (0.5) | 0 (0) | 2 (1.9) |
| Primary Cardiac Diagnosis | | | |
| Abnormal mixing | | | |
| Transposition of the great arteries (TGA) | - | - | 45 |
| Truncus arteriosus | - | - | 3 |
| Double outlet right ventricle | - | - | 3 |
| Total anomalous pulmonary venous drainage | - | - | 1 |
| Left sided heart lesions | | | |
| Aortic arch anomalies^a^ | - | - | 25 |
| Hypoplastic left heart syndrome(HLHS) | - | - | 7 |
| Aortic Stenosis | - | - | 3 |
| Right sided heart lesions | | | |
| Tetralogy of Fallot | - | - | 13 |
| Pulmonary atresia | - | - | 4 |
| Pulmonary stenosis | - | - | 2 |
| Tricuspid atresia | - | - | 1 |
| Neurodevelopmental follow-up assessments | | | |
| Number with follow-up assessment, n (%) | 345 (100) | 57 (78) | 65 of 88 infants eligible for follow-up (74)^b^ |
| Corrected age at assessment, median (IQR) | 18.4 (18.1-19.2) | 18.6 (18.2-19.3) | 22.29 (22.06-23.84) |
| Cognitive Composite Score, median (IQR) | 100 (95-110) | 100 (90-105) | 95 (85-100) |
| Motor Composite Score, median (IQR) | 103 (97-110) | 97 (91-107) | 94 (91-103) |
| Language Composite Score, median (IQR) | 100 (89-109) | 100 (86-112) | 91 (79-103) |
| Parents first language not English, n (%)^c^ | 152 (44.1) | 15 (26.3) | 15 (25) |
| ^a^Coarctation of the aorta n=22, hypoplastic aortic arch n=2 or interrupted aortic arch n=1; ^b^19 infants too young for follow-up; ^c^Missing data controls n=14; preterms n=3; CHD n=5 | | | |

| **Table S2**. Associations between demographic variables, allometric scaling and multivariate morphological features in typically developing infants | | | |
| --- | --- | --- | --- |
| **Log(SA)~Log(Supratentorial Volume)** | | | |
| Model | B (95% CI) | T | P (p_FWE_) |
| GA | 0.021 (-0.013- 0.054) | 1.20 | 0.225 (0.900) |
| PMA | -0.013 (-0.041- 0.015) | -0.906 | 0.357 (1.00) |
| PMA^2^ | <-0.001 (<-0.001- <0.001) | -0.916 | 0.366 (1.00) |
| Sex | -0.076 (-0.141- -0.010) | -2.26 | 0.029 (0.144) |
| Multiple birth | -0.069 (-0.244- 0.107) | -0.771 | 0.440 (1.00) |
| **Log(CT)~ log(Supratentorial Volume)** | | | |
| GA | -0.028 (-0.006- 0.004) | -1.74 | 0.086 (0.296) |
| PMA | -0.018 (-0.044- 0.008) | -1.32 | 0.190 (0.296) |
| PMA^2^ | 0.002 (<-0.001- 0.005) | 1.79 | 0.074 (0.296) |
| Sex | 0.054 (-0.008- 0.117) | 1.70 | 0.089 (0.296) |
| Multiple birth | 0.189 (0.024- 0.353) | 2.25 | 0.026 (0.130) |
| **Log(total SA*CT^1/2^) ~ log(outer SA)** | | | |
| GA | 0.014 (-0.031- 0.059) | 0.598 | 0.543 (1.00) |
| PMA | 0.028 (-0.011- 0.066) | 1.43 | 0.155 (0.620) |
| PMA^2^ | <0.001 (<-0.001- <0.001) | 1.53 | 0.119 (0.595) |
| Sex | 0.012 (-0.078- 0.102) | 0.264 | 0.786 (1.00) |
| Multiple birth | -0.038 (-0.265- 0.189) | -0.328 | 0.742 (1.00) |
| **Multivariate morphological offset term** | | | |
| GA | -0.003 (-0.006- <-0.001) | -2.66 | 0.010 (0.049)* |
| PMA | 0.001 (-0.062- 0.065) | 0.044 | 0.962 (1.00) |
| PMA^2^ | <0.001 (<-0.001- <0.001) | 0.509 | 0.609 (1.00) |
| Sex | -0.002 (-0.007- 0.002) | -1.06 | 0.295 (0.885) |
| Multiple birth | -0.014 (-0.027- <-0.001) | -2.11 | 0.036 (0.144) |
| **Multivariate morphological isometric term** | | | |
| GA | -0.037 (-0.048- -0.025) | -5.99 | <0.001 (<0.001)* |
| PMA | -0.195 (-0.512- 0.122) | -1.21 | 0.231 (0.302) |
| PMA^2^ | 0.003 (<-0.001- 0.007) | 1.79 | 0.075 (0.225) |
| Sex | 0.016 (-0.005- 0.038) | 1.47 | 0.151 (0.302) |
| Multiple birth | -0.090 (-0.155- -0.024) | -2.70 | 0.006 (0.024)* |
| **Multivariate morphological shape term** | | | |
| GA | 0.041 (0.020- 0.061) | 3.87 | <0.001 (<0.001)* |
| PMA | 0.540 (-0.006- 1.09) | 1.94 | 0.054 (0.164) |
| PMA^2^ | -0.007 (-0.013- <0.001) | -2.05 | 0.041 (0.164) |
| Sex | 0.020 (-0.017- 0.058) | 1.06 | 0.287 (0.574) |
| Multiple birth | 0.048 (-0.064- 0.162) | 0.851 | 0.381 (0.574) |
| * significant p_FWE_<0.05 | | | |

| **Table S3.** Associations between demographic variables, allometric scaling and multivariate morphological features in preterm infants and infants with CHD | | | |
| --- | --- | --- | --- |
| **Log(SA)~Log(Supratentorial Volume) preterms/CHD** | | | |
| Model | B (95% CI) | T | P (p_FWE_) |
| GA | -0.006 (-0.027- 0.016)/-0.011 (-0.083- 0.062) | -0.533/-0.299 | 0.592 (1.00)/0.759 (1.00) |
| PMA | -0.015 (-0.052- 0.022)/-0.025 (-0.090- 0.040) | -0.810/-0.754 | 0.421 (1.00)/0.441 (1.00) |
| PMA^2^ | <-0.001 (<-0.001- <0.001)/ <-0.001 (-0.001- <0.001) | -0.827/-0.796 | 0.415 (1.00)/0.430 (1.00) |
| Sex | -0.053 (-0.157- 0.052)/ -0.037 (-0.175- 0.100) | -1.01/-0.539 | 0.306 (1.00)/0.592 (1.00) |
| Multiple birth | 0.063 (-0.103- 0.229)/0.200 (-0.174- 0.574) | 0.761/1.06 | 0.454 (1.00)/0.279 (1.00) |
| **Log(CT)~ log(Supratentorial Volume)** | | | |
| GA | -0.004 (-0.027- 0.018)/-0.041 (-0.117- 0.034) | -0.379/-1.09 | 0.705 (1.00)/0.269 (1.00) |
| PMA | 0.009 (-0.030- 0.049)/-0.034 (-0.102-0.034) | 0.491/-1.00 | 0.633 (1.00)/0.324 (1.00) |
| PMA^2^ | <0.001 (<-0.001- <0.001)/<-0.001 (-0.001- <0.001) | 0.526/-0.974 | 0.592 (1.00)/0.332 (1.00) |
| Sex | 0.081 (-0.028- 0.190)/0.030 (-0.115-0.175) | 1.48/0.407 | 0.141 (1.00)/0.693 (1.00) |
| Multiple birth | 0.002 (-0.174- 0.178)/-0.049 (-0.444-0.346) | 0.023/-0.247 | 0.982 (1.00)/0.798 (1.00) |
| **Log(total SA*CT^1/2^) ~ log(outer SA)** | | | |
| GA | 0.006 (-0.020- 0.032)/-0.010 (-0.115- 0.095) | 0.536/-0.186 | 0.664 (1.00)/0.856 (1.00) |
| PMA | -0.006 (-0.053- 0.041)/-0.023 (-0.112- 0.067) | -0.265/-0.500 | 0.777 (1.00)/0.621 (1.00) |
| PMA^2^ | <-0.001 (<-0.001- <0.001)/ <-0.001 (-0.001- <0.001) | -0.257/-0.483 | 0.795 (1.00)/0.624 (1.00) |
| Sex | -0.040 (-0.169- 0.089)/-0.009 (-0.179- 0.161) | -0.618/-0.104 | 0.537 (1.00)/0.918 (1.00) |
| Multiple birth | 0.076 (-0.122- 0.275)/0.229 (-0.240- 0.698) | 0.767/0.968 | 0.442 (1.00)/0.333 (1.00) |
| **Multivariate morphological offset term** | | | |
| GA | 0.003 (0.002- 0.004)/-0.002 (-0.010- 0.005) | 4.10/-0.589 | <0.001 (0.006)*/0.553 (1.00) |
| PMA | 0.135 (0.041- 0.229)/0.101 (-6.24- 0.265) | 2.86/1.21 | 0.006 (0.156)/0.228 (1.00) |
| PMA^2^ | -0.001 (-0.003- <-0.001)/-0.001 (-0.003- 0.001) | -2.49/-0.979 | 0.016 (0.390)/0.330 (1.00) |
| Sex | 0.002 (-0.006- 0.009)/-0.002 (-0.008- 0.004) | 0.401/-0.722 | 0.701 (1.00)/0.473 (1.00) |
| Multiple birth | 0.002 (-0.006- 0.004)/-0.009 (-0.021- 0.003) | 0.498/-1.43 | 0.631 (1.00)/0.161 (1.00) |
| **Multivariate morphological isometric term** | | | |
| GA | 0.016 (0.007- 0.025)/-0.008 (-0.061- 0.044) | 3.39/-0.305 | 0.002 (0.043)*/0.770 (1.00) |
| PMA | 1.10 (0.486-1.71)/-0.164 (-1.31- 0.977) | 3.57/-0.285 | <0.001 (0.017)*/0.777 (1.00) |
| PMA^2^ | -0.012 (-0.020- -0.005)/0.003 (-0.011- 0.017) | -3.31/0.420 | <0.001 (0.012)*/0.675 (1.00) |
| Sex | 0.029 (-0.020- 0.079)/0.37 (-0.002- 0.076) | 1.20/1.85 | 0.233 (1.00)/0.067 (1.00) |
| Multiple birth | 0.068 (0.003- 0.133)/ -0.067 (-0.154- 0.019) | 2.09/-1.55 | 0.037 (1.00)/0.132 (1.00) |
| **Multivariate morphological shape term** | | | |
| GA | 0.014 (<-0.001- 0.029)/-0.060 (-0.153- 0.034) | 1.89/-1.26 | 0.062 (1.00)/0.209 (1.00) |
| PMA | 0.028 (-0.957- 1.01)/1.32 (-0.716- 3.36) | 0.056/1.29 | 0.960 (1.00)/0.200 (1.00) |
| PMA^2^ | <-0.001 (-0.012- 0.011)/-0.015 (-0.041- 0.010) | -0.075/-1.19 | 0.945 (1.00)/0.233 (1.00) |
| Sex | 0.077 (-0.002- 0.156)/0.098 (0.028- 0.168) | 1.95/2.76 | 0.051 (1.00)/0.007 (0.210) |
| Multiple birth | -0.006 (-0.110- 0.098)/0.057 (-0.097- 0.211) | -0.112/0.739 | 0.919 (1.00)/0.441 (1.00) |
| * significant p_FWE_<0.05 | | | |

| **Table S4.** Associations between scaling relationship and multivariate morphological term deviance z-scores and cognitive language and motor abilities in typically developing infants | | | | | | |
| --- | --- | --- | --- | --- | --- | --- |
| Composite score | B | 2.5% CI bound | 97.5% CI bound | t | p | p_FWE_ |
| **total surface area*cortical thickness^1/2^** | | | | | | |
| cognition | 0.180 | -0.942 | 1.302 | 0.315 | 0.751 | 1.00 |
| language | -0.210 | -1.781 | 1.362 | -0.262 | 0.799 | 1.00 |
| motor | 0.404 | -0.612 | 1.419 | 0.782 | 0.423 | 1.00 |
| **multivariate isometric term** | | | | | | |
| cognition | 0.332 | -0.795 | 1.458 | 0.579 | 0.564 | 1.00 |
| language | -0.011 | -1.618 | 1.596 | -0.013 | 0.989 | 1.00 |
| motor | -0.389 | -1.409 | 0.630 | -0.751 | 0.452 | 1.00 |
| **multivariate offset term** | | | | | | |
| cognition | -0.002 | -1.124 | 1.121 | -0.003 | 0.997 | 1.00 |
| language | -0.336 | -1.915 | 1.243 | -0.418 | 0.678 | 1.00 |
| motor | 0.035 | -0.982 | 1.051 | 0.067 | 0.947 | 1.00 |
| t**otal surface area** | | | | | | |
| cognition | 0.649 | -0.470 | 1.768 | 1.141 | 0.248 | 1.00 |
| language | 0.330 | -1.247 | 1.908 | 0.412 | 0.676 | 1.00 |
| motor | 0.515 | -0.499 | 1.529 | 0.999 | 0.321 | 1.00 |
| **multivariate shape term** | | | | | | |
| cognition | 0.651 | -0.467 | 1.770 | 1.145 | 0.255 | 1.00 |
| language | 0.303 | -1.282 | 1.888 | 0.376 | 0.699 | 1.00 |
| motor | 0.581 | -0.432 | 1.594 | 1.128 | 0.265 | 1.00 |
| **mean cortical thickness** | | | | | | |
| cognition | -0.428 | -1.548 | 0.693 | -0.751 | 0.456 | 1.00 |
| language | -0.239 | -1.827 | 1.348 | -0.297 | 0.761 | 1.00 |
| motor | -0.569 | -1.582 | 0.445 | -1.103 | 0.257 | 1.00 |
| *significant p_FWE_<0.05; adjusting for index of multiple deprivation and for language, if a parent speaks English as a second language | | | | | | |

| **Table S5.** Whole brain cortical metrics across all groups | | | |
| --- | --- | --- | --- |
| Metric | Controls | Preterms | CHD |
| Total surface area (cm^2^) | 974 (871-108) | 900 (816- 105) | 792 (721-845) |
| Mean cortical thickness (mm) | 1.11 (1.07-1.14) | 1.14 (1.10- 1.17) | 1.08 (1.06-1.10) |
| Gyrification index (total surface area/exposed surface area) | 2.62 (2.47-2.73) | 2.44 (2.33- 2.60) | 2.37 (2.27-2.46) |
| Supratentorial volume (cm^3^) | 335 (309-368) | 325 (300- 364) | 280 (262-298) |
| **Effect of group in typically developing controls (reference) and preterm infants** | | | |
|  | B (95% CI) | t-score | p (p_FWE_) |
| Total surface area | -5123 (-7482- -2764) | -4.27 | <0.001 (<0.001)* |
| Mean cortical thickness | 0.025 (0.014-0.037) | 4.28 | <0.001 (<0.001)* |
| Gyrification index | -0.143 (-0.176- -0.111) | -8.65 | <0.001 (<0.001)* |
| Supratentorial volume | -117 (-199- -33.9) | -2.77 | 0.007 (0.007)* |
| **Effect of group in typically developing controls (reference) and infants with CHD** | | | |
| Total surface area | -4251 (-6229- -2274) | -4.23 | <0.001 (<0.001)* |
| Mean cortical thickness | 0.005 (-0.008- 0.013) | 0.471 | 0.649 (0.649) |
| Gyrification index | -0.044 (-0.076- -0.011) | -2.64 | 0.010 (0.020)* |
| Supratentorial volume | -195 (-268- -121) | -5.23 | <0.001 (<0.001)* |
| **Effect of group in preterm infants (reference) and infants with CHD** | | | |
| Total Surface Area | 135 (-155- 424) | 0.917 | 0.370 (0.740) |
| Mean Cortical Thickness | -0.019 (-0.034- -0.003) | -2.30 | 0.022 (0.066) |
| Gyrification index | 0.105 (0.066-0.144) | 5.32 | <0.001 (<0.001)* |
| Supratentorial volume | -495 (-1605- 618) | -0.877 | 0.397 (0.740) |
| *significant p_FWE_<0.05; adjusting for PMA, PMA^2^, sex and multiple birth | | | |

| **Table S6.** Associations between cortical metrics and neurodevelopmental outcomes in typically developing controls, preterm infants and CHD | | | | | | |
| --- | --- | --- | --- | --- | --- | --- |
| Metric | B | 2.5% CI bound | 97.5% CI bound | t-score | p | p_FWE_ |
| **Cognitive composite score** | | | | | | |
| *Typically developing controls* | | | | | | |
| Mean cortical thickness | -6.51 | -32.3 | 19.3 | -0.497 | 0.614 | 1.00 |
| Supratentorial brain volume | <0.001 | <0.001 | <0.001 | 1.41 | 0.149 | 1.00 |
| Total surface area | <0.001 | <0.001 | <0.001 | 1.79 | 0.073 | 0.876 |
| Gyrification index | 4.99 | -4.00 | 13.97 | 1.09 | 0.268 | 1.00 |
| *Preterm-born infants* | | | | | | |
| Mean cortical thickness | -69.3 | -128 | -10.8 | -2.33 | 0.021 | 0.231 |
| Supratentorial brain volume | <0.001 | <0.001 | <0.001 | 0.099 | 0.923 | 1.00 |
| Total surface area | <-0.001 | <-0.001 | <0.001 | -0.007 | 0.995 | 1.00 |
| Gyrification index | 7.38 | -10.8 | 25.6 | 0.796 | 0.424 | 1.00 |
| *Infants with CHD* | | | | | | |
| Mean cortical thickness | 45.3 | -18.0 | 108 | 1.41 | 0.160 | 1.00 |
| Supratentorial brain volume | <-0.001 | <-0.001 | <0.001 | -0.508 | 0.606 | 1.00 |
| Total surface area | <-0.001 | <-0.001 | <0.001 | -0.234 | 0.812 | 1.00 |
| Gyrification index | 0.384 | -18.9 | 19.7 | 0.039 | 0.964 | 1.00 |
| **Language composite score** | | | | | | |
| *Typically developing controls* | | | | | | |
| Mean cortical thickness | -6.41 | -42.8 | 29.9 | -0.347 | 0.731 | 1.00 |
| Supratentorial brain volume | <0.001 | <0.001 | <0.001 | 0.308 | 0.766 | 1.00 |
| Total surface area | <0.001 | <0.001 | <0.001 | 0.473 | 0.630 | 1.00 |
| Gyrification index | 0.502 | -12.0 | 13.0 | 0.079 | 0.935 | 1.00 |
| *Preterm infants* | | | | | | |
| Mean cortical thickness | -114 | -201 | -27.1 | -2.58 | 0.009 | 0.103 |
| Supratentorial brain volume | <-0.001 | <-0.001 | <0.001 | -0.069 | 0.942 | 1.00 |
| Total surface area | <-0.001 | <-0.001 | <0.001 | -0.001 | 0.999 | 1.00 |
| Gyrification index | 15.1 | -10.7 | 40.9 | 1.15 | 0.239 | 1.00 |
| *Infants with CHD* | | | | | | |
| Mean cortical thickness | 55.9 | -42.6 | 154 | 1.12 | 0.263 | 1.00 |
| Supratentorial brain volume | <-0.001 | <-0.001 | <0.001 | -0.042 | 0.974 | 1.00 |
| Total surface area | <0.001 | <0.001 | <0.001 | 0.354 | 0.723 | 1.00 |
| Gyrification index | 9.83 | -19.0 | 38.7 | 0.670 | 0.506 | 1.00 |
| **Motor composite scores** | | | | | | |
| *Typically developing controls* | | | | | | |
| Mean cortical thickness | -13.9 | -37.5 | 9.72 | -1.16 | 0.246 | 1.00 |
| Supratentorial brain volume | <-0.001 | <-0.001 | <0.001 | -0.024 | 0.984 | 1.00 |
| Total surface area | <0.001 | <0.001 | <0.001 | 0.437 | 0.654 | 1.00 |
| Gyrification index | 4.30 | -3.95 | 12.55 | 1.026 | 0.288 | 1.00 |
| *Preterm infants* | | | | | | |
| Mean cortical thickness | <0.001 | <0.001 | <0.001 | 0.471 | 0.627 | 1.00 |
| Supratentorial brain volume | -6.92 | -61.1 | 47.3 | -0.251 | 0.802 | 1.00 |
| Total surface area | <0.001 | <0.001 | <0.001 | 0.418 | 0.672 | 1.00 |
| Gyrification index | 9.85 | -6.92 | 26.6 | 1.15 | 0.245 | 1.00 |
| *Infants with CHD* | | | | | | |
| Mean cortical thickness | 35.3 | -23.5 | 94.1 | 1.18 | 0.229 | 1.00 |
| Supratentorial brain volume | <-0.001 | <-0.001 | <0.001 | -0.149 | 0.891 | 1.00 |
| Total surface area | <-0.001 | <-0.001 | <0.001 | -0.297 | 0.761 | 1.00 |
| Gyrification index | -4.66 | -22.5 | 13.2 | -0.514 | 0.608 | 1.00 |
| *significant p_FWE_<0.05; linear regression with permutation testing (n=5000) adjusting for index of multiple deprivation, sex, PMA, PMA^2^ and multiple birth | | | | | | |

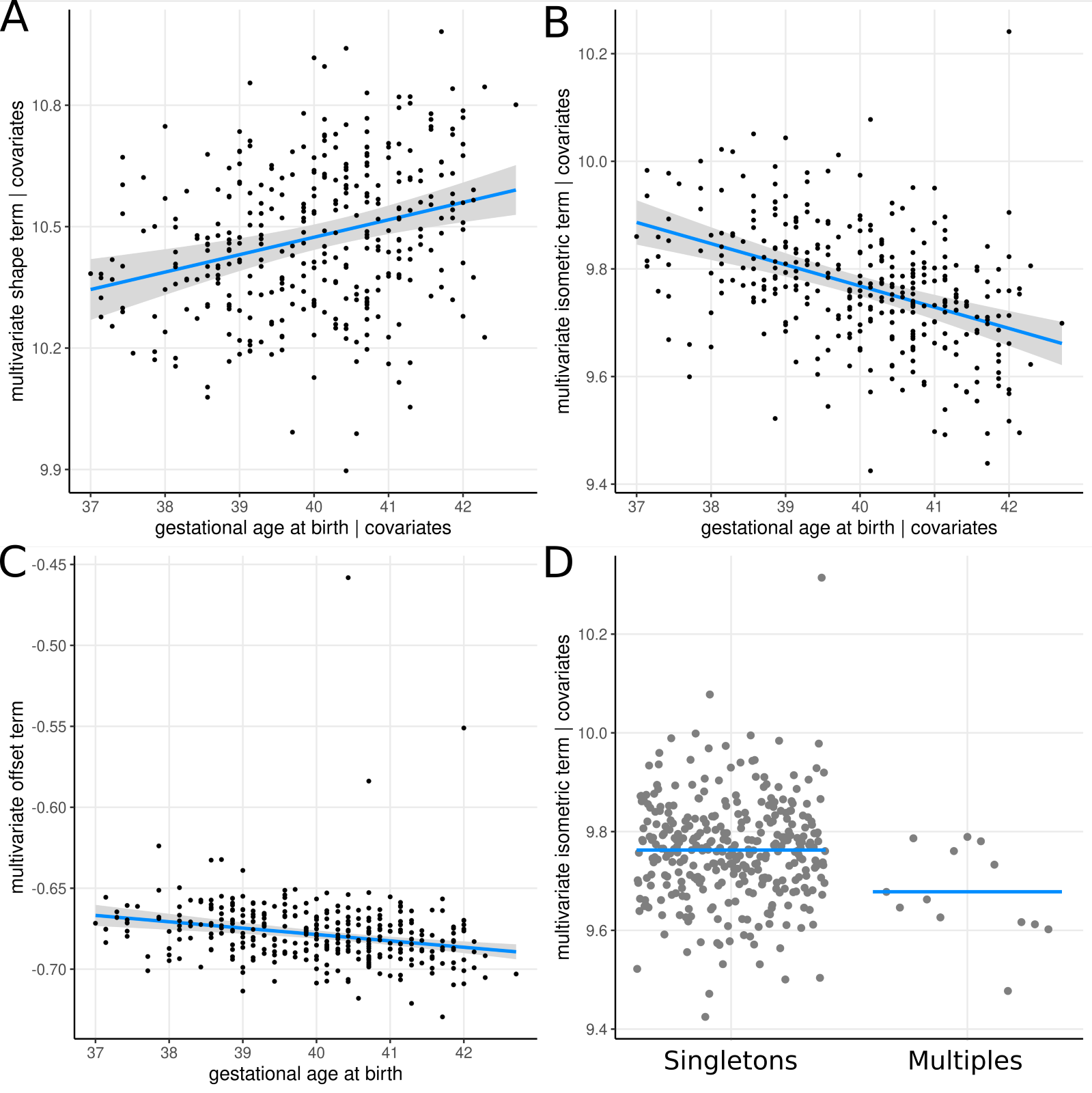

**Figure S1.** A-C) associations between gestational age at birth and multivariate morphological terms in typically developing control infants, adjusting for PMA, PMA^2^ multiple birth and sex. D) Associations between multiple birth and multivariate isometric term adjusting for adjusting for GA, PMA, PMA^2^ and sex
